# Hemodynamics with Mechanical Circulatory Support Devices Using a Cardiogenic Shock Model

**DOI:** 10.1101/2023.08.16.23294188

**Authors:** Kazuyuki Yahagi, Gohki Nishimura, Kei Kuramoto, Yusuke Tsuboko, Kiyotaka Iwasaki

**Affiliations:** Cooperative Major in Advanced Biomedical Sciences, Joint Graduate School of Tokyo Women’s Medical University and Waseda University, Waseda University, Tokyo, Japan; Division of Cardiology, Mitsui Memorial Hospital, Tokyo, Japan; Department of Modern Mechanical Engineering, School of Creative Science and Engineering, Waseda University, Tokyo, Japan; Waseda Research Institute for Science and Engineering, Waseda University, Tokyo, Japan; Department of Integrative Bioscience and Biomedical Engineering, Graduate School of Advanced Science and Engineering, Waseda University, Tokyo, Japan; Institute for Medical Regulatory Science, Comprehensive Research Organization, Waseda University, Shinjuku, Tokyo, Japan

**Keywords:** Cardiogenic shock, Impella, Mechanical circulatory support, Veno-arterial extracorporeal membrane oxygenation

## Abstract

**Background:** Mechanical circulatory support (MCS) devices, including veno-arterial extracorporeal membrane oxygenation (VA-ECMO) and Impella, have been widely used for patients with cardiogenic shock (CS). However, hemodynamics with each device and combination therapy is not thoroughly understood. We aimed to elucidate the hemodynamics with MCS using a CS pulsatile flow model.

**Methods and Results:** A pulsatile flow circulation system comprising a ventricle, aortic valve, mitral valve, elastic artery, and venous reservoir was developed. Hemodynamics with Impella CP, VA-ECMO, and a combination of Impella CP and VA-ECMO were assessed based on the pressure and flow under support with each device and the pressure-volume loop of the ventricle model. The self-recirculation with Impella was assessed using particle image velocimetry. The Impella CP device with CS status resulted in increased afterload, leading to an increase in aortic pressure and a decrease in end-diastolic volume and end-diastolic pressure (EDP). VA-ECMO support resulted in increased afterload, leading to a significant increase in aortic pressure with an increase in end-systolic volume and EDP and decreasing venous reservoir pressure. The combination of Impella CP and VA-ECMO led to left ventricular unloading, regardless of increase in afterload. Self-recirculation with Impella CP was observed in a patient with significant aortic insufficiency, leading to reduced hemodynamic support.

**Conclusions:** The Impella system improved hemodynamics with ventricular unloading. Hemodynamic support with Impella and VA-ECMO should be a promising combination for patients with severe CS. Physicians should be alert to the presence of significant aortic insufficiency, leading to self-recirculation with Impella.

**Clinical Perspective:** *What is new?:*

- This is a non-clinical study using a novel experimental pulsatile flow system with venous function that can be used to assess MCS, including Impella and VA-ECMO.
- The self-recirculation phenomenon of the Impella device was assessed by a flow visualization method using particle imaging velocimetry.

*What are the clinical implications?:*

- The combination of Impella and VA-ECMO led to left ventricular unloading, regardless of the increase in afterload.
- VA-ECMO could increase arterial pressure owing to increased LV afterload and alter the direction of the aortic flow (antegrade or retrograde) depending on the flow level with VA-ECMO.
- Effective Impella flow was reduced during ventricular diastole when aortic insufficiency was present owing to self-recirculation with the Impella device.

## Introduction

Cardiogenic shock (CS) remains a clinical challenge associated with high mortality rates (1,2). Mechanical circulatory support (MCS) devices have been widely used for patients with CS, including those with advanced heart failure secondary to ischemic heart disease, cardiomyopathy, or myocarditis. ^1, 2^ Previously, intra-aortic balloon pumping (IABP) and veno-arterial extracorporeal membrane oxygenation (VA-ECMO) were the mainstay for the management of CS. ^3–5^ In 2012, Thiele et al. reported the results of the IABP-SHOCK II trial, which failed to show that mechanical support with IABP improved the outcomes of patients with acute myocardial infarction (AMI) with CS. ^6^ Moreover, there was no difference in all-cause mortality between IABP and control groups at the 6-year long-term follow-up. ^7^ Based on this randomized clinical trial, the current guidelines do not recommend the routine use of IABP therapy for CS. ^4, 8^ VA-ECMO is a rescue therapy that involves powerful hemodynamic support for patients with severe CS. However, the disadvantage of VA-ECMO is an elevated left ventricular (LV) afterload, resulting in greater LV workload. Impella, a catheter with a small built-in axial flow pump (Abiomed, Danvers, MA, USA), has been approved for use in CS. Impella is expected to have an unloading effect because it can pump blood directly from the left ventricle. ^9^ Additionally, the combination of Impella and VA-ECMO improves hemodynamics and mortality. ^10, 11^ However, the mortality rate of patients with refractory shock is extremely high. ^12^ Hemodynamics with Impella or the combination of Impella and VA-ECMO has not been thoroughly understood.

Assessment of hemodynamics, including pressure and flow in the aorta and venous system, and ventricular function, which is represented using a pressure-volume loop during a complete cardiac cycle, is valuable for understanding MCS. ^13, 14^ Although an in vitro circulation study is a powerful tool for understanding hemodynamics in a specific target, ^15, 16^ there has been no comprehensive study on hemodynamics with MCS. Here, we assessed the effects of Impella, VA-ECMO, and their combined use on hemodynamic changes using a clinically relevant circulatory system simulating CS.

## Methods

### Data Availability

The data that support the findings of this study are available from the corresponding author upon reasonable request.

### CS model with a pulsatile flow system

The pulsatile circulatory system consisted of a ventricular model, a mitral valve, an aortic valve, an elastic artery model, a systemic vascular resistance, and a venous reservoir with an elastic vessel model, which was modified based on our previous circulatory system (Figure 1A and 1B). ^16^ The silicone ventricular model was driven using a pneumatic console. The aortic valve model comprised a three-dimensional (3D)-printed polymer frame with a tri-leaflet bovine pericardial tissue cusp (Figure 1C). The mitral valve model comprised a 3D-printed polymer frame with a bileaflet polyurethane valve. ^17^ The artery model was made of silicone and included an aorta, a branch of the aorta (brachiocephalic artery), and the iliac arteries. The dimensions of these models were determined based on human data. ^18–21^ Digital Imaging and COmmunications in Medicine images were imported into segmentation software (Mimics, Materialize, Leuven, Belgium), and the 3D models were reconstructed. The total volume capacity of the circulation loop was approximately 5500 ml, including 500 ml in the ventricle, 1500 ml on the arterial side, and 3500 ml on the venous side. The mean circulatory filling pressure was controlled by the volume of physiological saline solution and was set at 7 mmHg for the normal condition and 13 mmHg for the CS condition.

**Figure 1:**
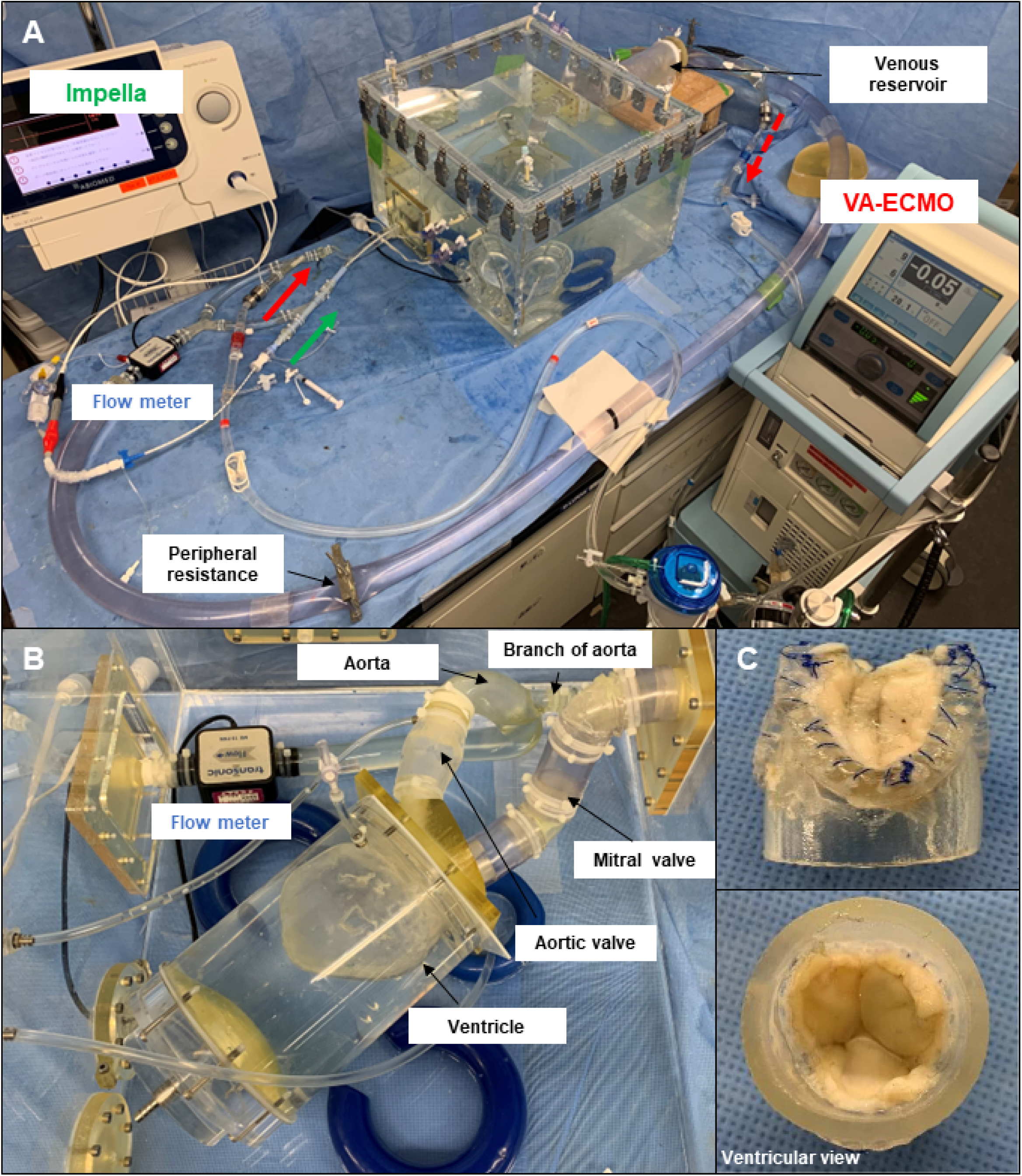
Experimental setup. (A) A pulsatile circulation model with a venous system; (B) A ventricular model with an aorta and a branch (brachiocephalic artery), an aortic valve, and a mitral valve; (C) An aortic valve made of bovine pericardium.

### Assessment of hemodynamics

Heart rate was maintained at 75 beats/min at a fixed rate. Pressure was measured in the ventricle, aorta, and venous reservoir using pressure transducers (PXMK10200; Edwards Lifesciences, Irvine, CA, USA). The flow was measured at the aorta, distal artery (iliac artery), and branch of the aorta (brachiocephalic artery) using an ultrasonic flow sensor (ME-PXN ME19PXN325; Transonic, Ithaca, NY, USA). The flow sensor in the distal artery was located at the distal part of the cannulation site for the VA-ECMO outlet (Figure 1A). The mean total flow was calculated as the mean flow at the distal artery (iliac artery) and brachiocephalic artery. Aortic pressure was adjusted by peripheral resistance with a tube clamp. Peripheral resistance in the CS condition was the same as that in the normal condition. The brachiocephalic artery flow was adjusted using a tube clamp. Previous reports have shown that normal cerebral blood flow represents approximately 14% of the total cardiac output, which reduces in heart failure. ^22^ A conductance catheter (CD Leycom, Zoetermeer, The Netherlands) was placed in the LV, and the pressure-volume loop of the LV, including mean peak ventricle pressure (VP), mean end-diastolic pressure (EDP), mean end-diastolic volume (EDV), and mean end-systolic volume (ESV), was assessed. Mean stroke volume (SV) was calculated as the difference between EDV and ESV. These parameters were measured under each set of experimental conditions.

### Assessment of self-recirculation

The self-recirculation phenomenon of the Impella device was assessed by a flow visualization method using particle imaging velocimetry (PIV). The pulsatile flow circulation system was modified for PIV assessment. To assess the influence of the severity of aortic insufficiency on the self-recirculation with Impella, two types of aortic valves were prepared. One was a soft silicone valve demonstrating without significant aortic regurgitation, and the other was a hard silicone valve demonstrating with aortic regurgitation.

A water glycerol solution mixed with fluorescent particles with a mean particle size of 13 μm and density of 1.1 g/cm^3^ (Fluostar, EBM, Tokyo, Japan) was circulated. The refractive index of the fluid was adjusted to 1.4050 to adjust that of the silicone, ensuring precise flow visualization. A pulsed Nd:YAG laser (DS30-527; Photonics Industries, Ronkonkoma, NY, USA) was used to emit circulating fluorescent particles, and the images were captured using a high-speed camera (Fastcam Mini WX50; Photoron, Tokyo, Japan). Image acquisition and processing were performed using imaging software (DaVis 10.2; LaVision, Göttingen, Germany).

Self-recirculation with the Impella device was defined as retrograde flow toward the inlet through the aortic valve because it was difficult to distinguish self-recirculation from native aortic regurgitant flow. Self-recirculation was assessed based on the total aortic regurgitant flow (TARF) in the proximal aortic valve. TARF was assessed based on regurgitant fraction by PIV analysis that was referred to as the regurgitant flow volume divided by the forward flow volume and is expressed in percent (%).

### CS model with MCS devices

Hemodynamics was assessed based on the CS status using MCS devices, including Impella CP and VA-ECMO (Senko Medical Instrument, Tokyo, Japan). Impella CP can deliver up to a mean maximum of 3.7 liters per minute (L/min) from the ventricle into the aorta. Impella CP has 10 performance levels ranging from P0 to P9 (0 to 46000 revolutions per minute [rpm]). In the circulation system, Impella CP was placed from the right iliac artery to the ventricle.VA-ECMO drained from the venous reservoir and returned to the left iliac artery.

## Results

### Hemodynamics of the normal and CS conditions

Under the normal condition, aortic pressure was 111/67 (88) mmHg, mean aortic flow was 4.3 L/min, mean flow at the brachiocephalic artery was 0.6 L/min (12.2% of total cardiac output), and mean venous reservoir pressure was 8.1 mmHg (Figure 2A). Regarding the CS status, aortic pressure was 75/35 (54) mmHg, mean aortic flow was 2.8 L/min, mean flow at the brachiocephalic artery was 0.4 L/min (12.5% of total cardiac output), and mean venous reservoir pressure was 13.1 mmHg. The pressure-volume loop is shown in Figure 2B. Under the normal condition, the peak VP, EDP, ESV, EDV, and SV were 137 mmHg, 13 mmHg, 107 ml, 169 ml, and 62 ml, respectively. Under the CS condition, a lower peak VP (88 mmHg), higher EDP (24 mmHg), higher ESV and EDV (148 and 199 ml, respectively), and smaller SV (51 ml) were observed compared with those under the normal conditions. Under the CS condition, a decrease in the end-systolic LV elastance (Ees) resulting from a decrease in contractility and the effective arterial elastance (Ea) moved to the right side due to increased preload (increased EDV) compared with those under the normal cardiac condition, which are well-known changes for low cardiac output (13). The pressure-volume loop was reproduced in accordance with a previous report (13). Using a clinically relevant CS model, we further assessed the influence of MCS on hemodynamics.

**Figure 2:**
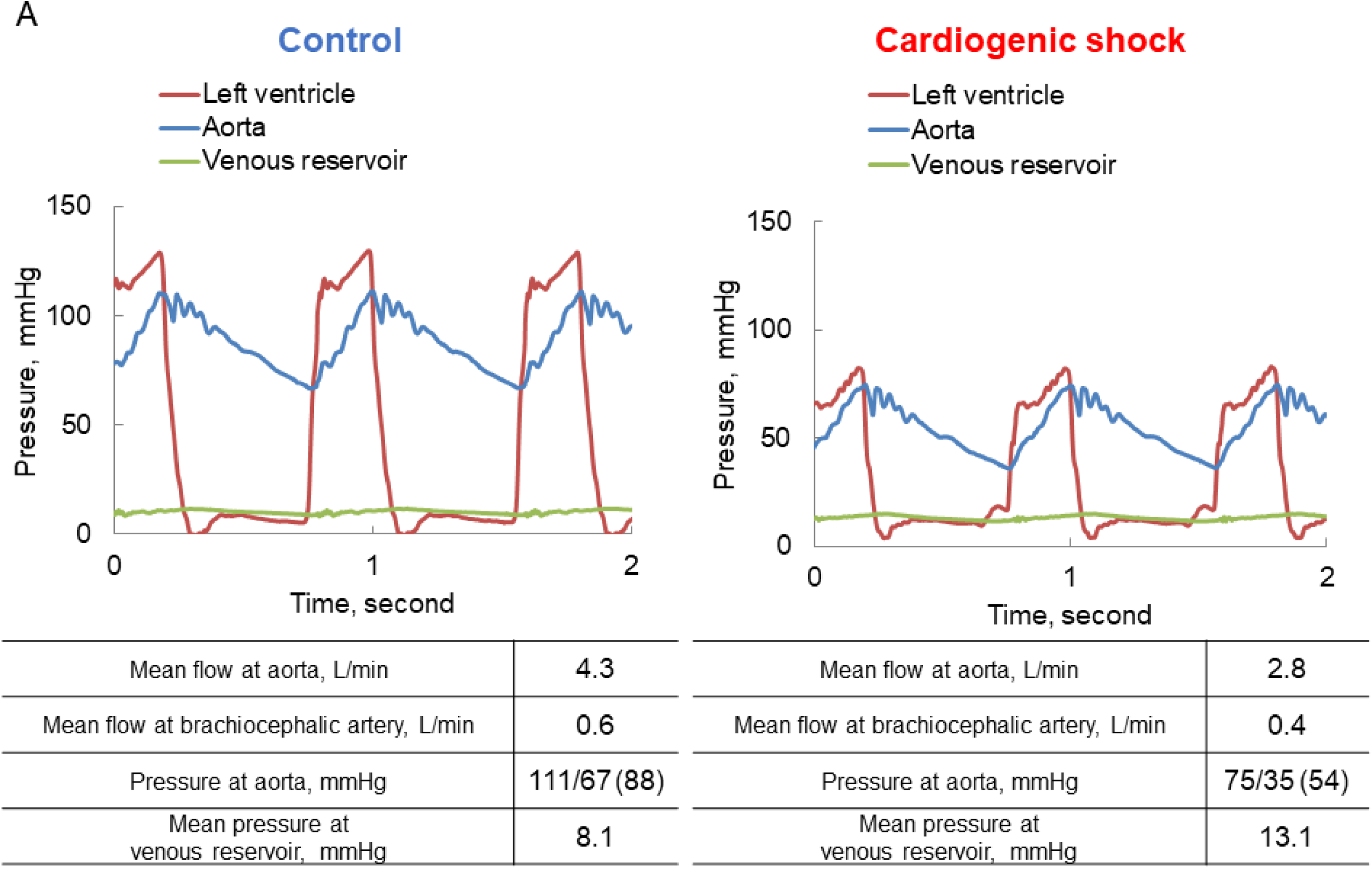
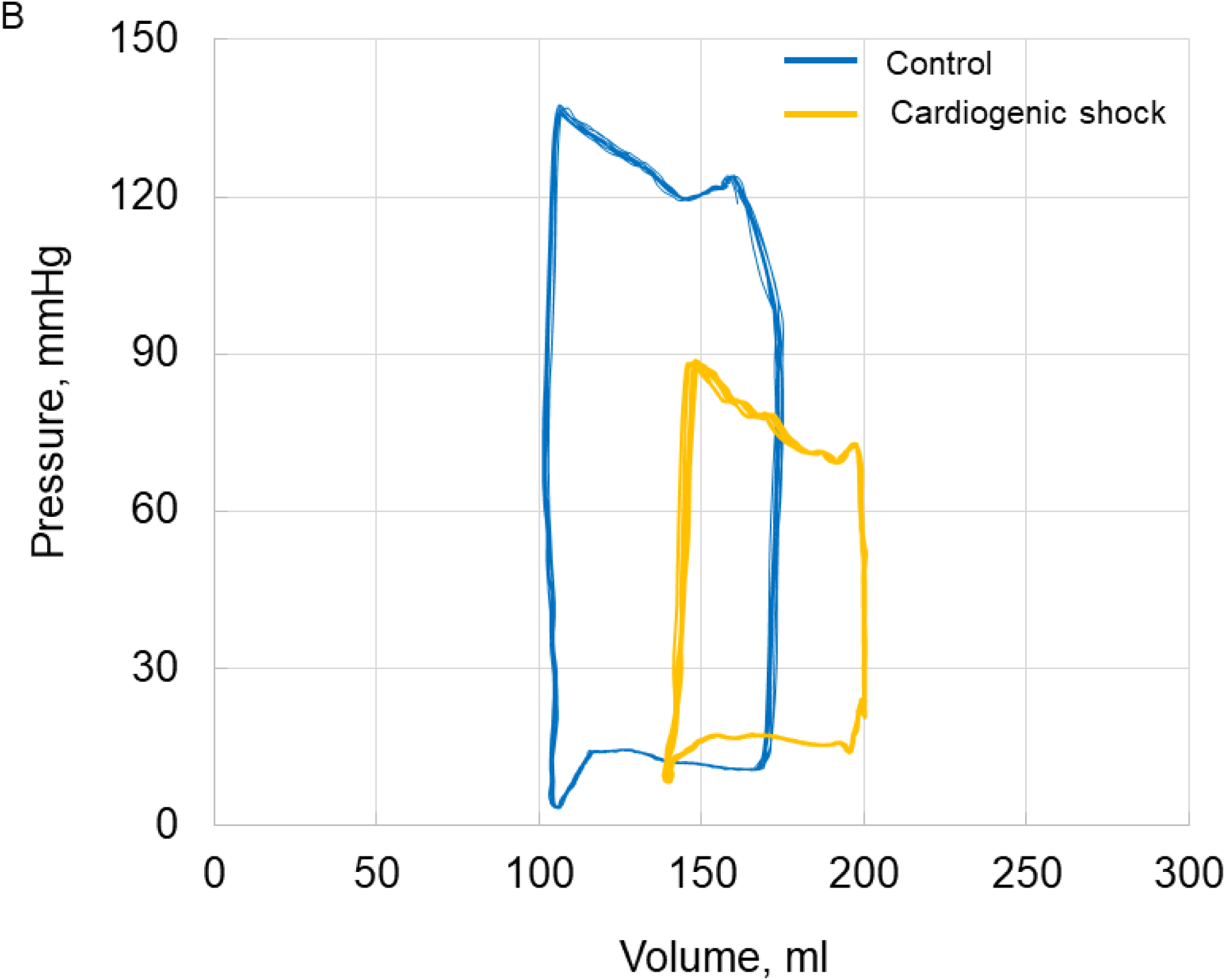
Representative hemodynamic waveform and pressure-volume loop. Representative hemodynamic waveform (A) and pressure-volume loop (B) of the left ventricle of control and cardiogenic shock model. Blue line, control; Yellow line, cardiogenic shock.

### Changes in CS hemodynamics by MCS

The flow level of Impella CP increased stepwise from P1 (23000 rpm) to P9 (46000 rpm), leading to increased mean total flow and mean aortic pressure (mean total flow: 2.70 L/min at P0 to 4.18 L/min at P9, mean pressure at the aorta: 46 mmHg at P0 to 76 mmHg at P9, respectively) (Table 1). Venous reservoir pressure decreased according to increased Impella flow (15.0 mmHg at P0 to 9.1 mmHg at P9). Based on the assessment of the pressure-volume loop, EDP/EDV decreased with increasing Impella flow level (Table 1, Figure 3A). Consequently, SV decreased by increasing Impella flow, which was driven by reduction in EDV.

**Figure 3:**
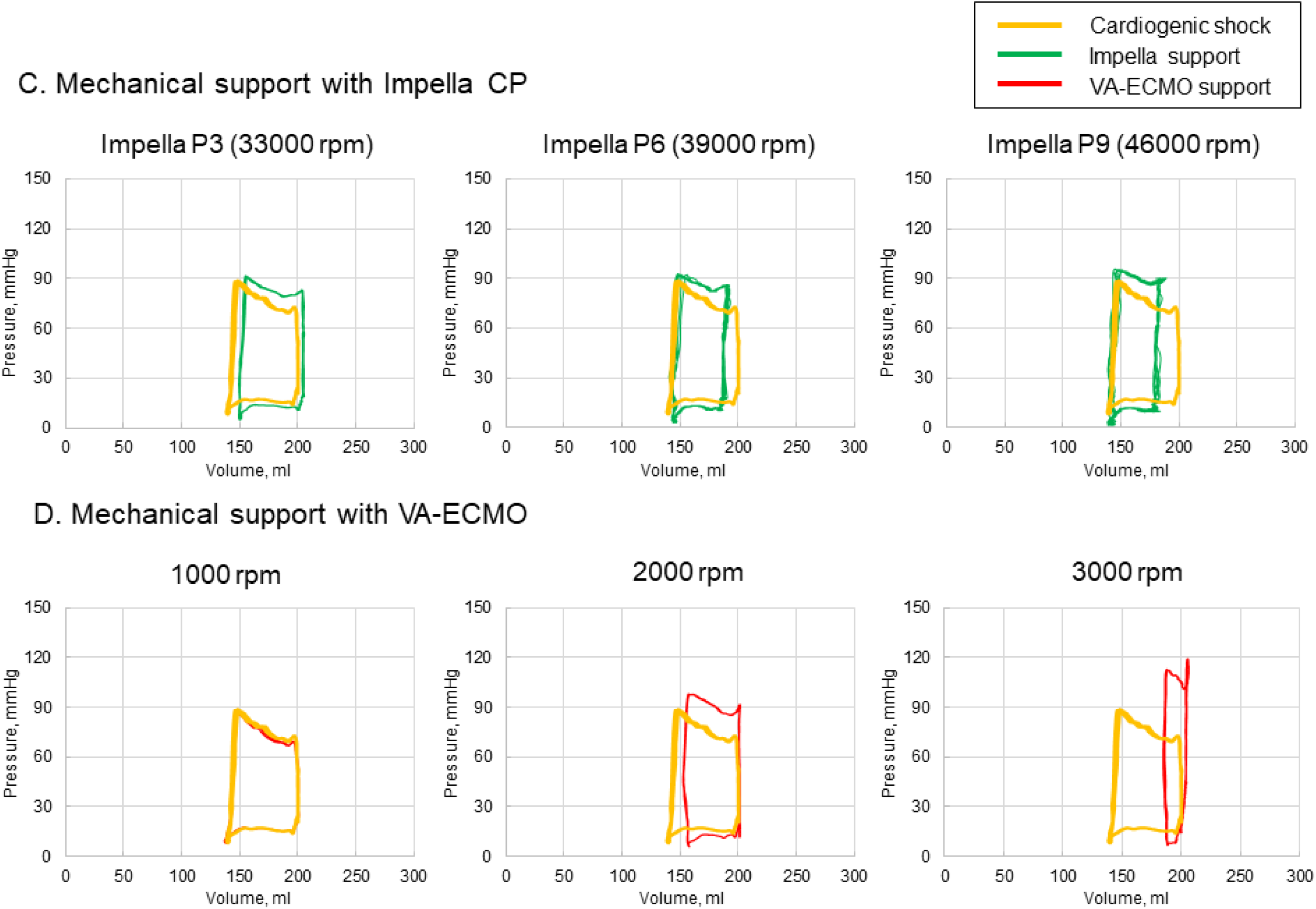
Representative pressure-volume loop of the left ventricle in the cardiogenic shock model. (A) Mechanical support with Impella CP at P3 (33000 rpm), P6 (39000 rpm), and P9 (46000 rpm); (B) Mechanical support with veno-arterial extracorporeal membrane oxygenation (VA-ECMO) at 1000, 2000, and 3000 rpm. Yellow line: cardiogenic shock; green line: Impella support; red line: VA-ECMO support.

**Table 1.** Hemodynamics with Impella CP support.

|  | Impella CP |  |  |  |  |  |  |  |  |  |
| --- | --- | --- | --- | --- | --- | --- | --- | --- | --- | --- |
|  | P0<br>0 rpm | P1<br>23<br>000<br>rpm | P2<br>31<br>000<br>rpm | P3<br>33<br>000<br>rpm | P4<br>35<br>000<br>rpm | P5<br>37<br>000<br>rpm | P6<br>39<br>000<br>rpm | P7<br>42<br>000<br>rpm | P8<br>44<br>000<br>rpm | P9<br>46<br>000<br>rpm |
| Mean total flow, L/min | 2.70 | 3.27 | 3.64 | 3.70 | 3.82 | 3.89 | 3.96 | 4.07 | 4.14 | 4.18 |
| Pressure at A, s/d (m), mmHg | 73/23 (46) | 78/39 (56) | 82/48 (64) | 83/52 (66) | 84/55 (68) | 85/57 (69) | 86/59 (71) | 88/61 (73) | 90/63 (74) | 91/65 (76) |
| Mean pressure at VR, mmHg | 15.0 | 12.3 | 11.3 | 10.9 | 10.5 | 10.4 | 10.0 | 9.4 | 9.3 | 9.1 |
| Peak LVP, mmHg | 88 | 87 | 90 | 91 | 92 | 92 | 92 | 94 | 95 | 95 |
| EDP, mmHg | 24 | 21 | 20 | 20 | 18 | 18 | 18 | 17 | 18 | 17 |
| ESV, ml | 148 | 157 | 155 | 156 | 156 | 154 | 150 | 147 | 147 | 148 |
| EDV, ml | 199 | 210 | 204 | 199 | 203 | 193 | 188 | 187 | 184 | 180 |
| Stroke volume, ml | 51 | 53 | 50 | 42 | 47 | 39 | 38 | 40 | 37 | 32 |

Mean total flow= mean flow of the iliac artery plus that of brachiocephalic trunk; A, aorta; EDP, end-diastolic pressure; EDV, end-diastolic volume; ESV, end-systolic volume; VR, venous reservoir; LVP, left ventricular pressure; s, systolic; d, diastolic; m, mean.

The flow level of VA-ECMO increased stepwise to 1000, 1500, 2000, 2500, and 3000 rpm, leading to increased mean total flow and mean aortic pressure (mean total flow: 2.70 L/min at 0 rpm to 4.47 L/min at 3000 rpm, mean pressure at the aorta: 46 mmHg at 0 rpm to 97 mmHg at 3000 rpm, respectively) (Table 2, Figure 3B). Venous reservoir pressure decreased according to increased VA-ECMO support (15.0 mmHg at 0 rpm to 7.0 mmHg at 3000 rpm). The pressure-volume loop became increasingly narrow (decreased SV) and taller (increased afterload), leading to an increased pressure-volume area (myocardial oxygen demand) with increasing VA-ECMO support. SV decreased with increasing VA-ECMO support, driven by an increase in ESV.

**Table 2.**
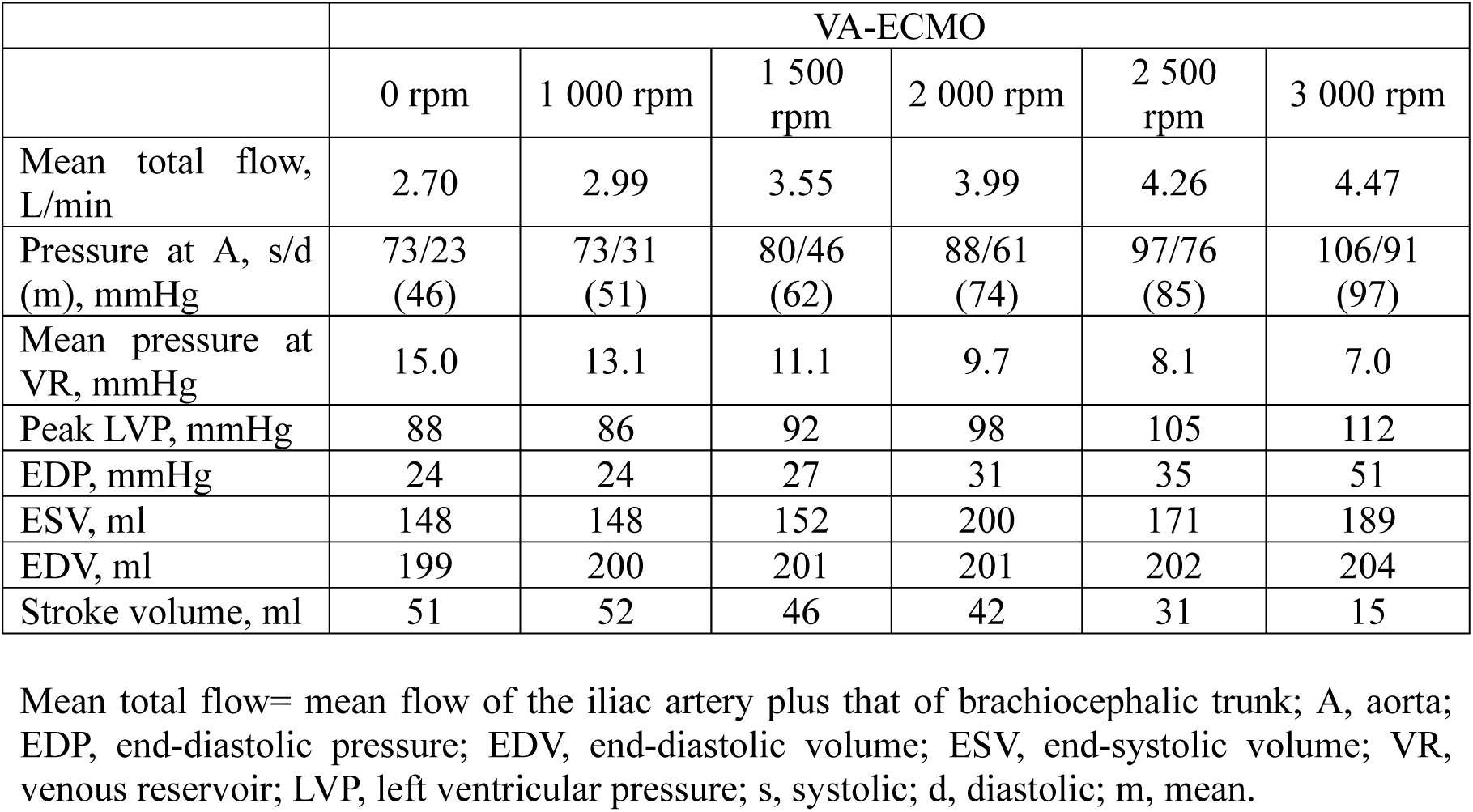
Hemodynamic parameters with VA-ECMO support.

Hemodynamics of the combination with Impella and VA-ECMO is shown in Table 3, and representative pressure-volume loops are shown in Figure 4. The mean total flow and mean aortic pressure increased, and the mean venous reservoir pressure decreased by increasing either Impella CP or VA-ECMO support. The combination of Impella CP (P9, 46000 rpm) and VA-ECMO (3000 rpm) showed maximum mean total flow (5.24 L/min), maximum mean arterial pressure (116 mmHg), and minimum venous reservoir pressure (5 mmHg). EDP decreased with increasing Impella flow level, even with VA-ECMO support. However, EDP was higher with high VA-ECMO support (3000 rpm) than with low VA-ECMO support (2000 rpm) due to increased afterload by VA-ECMO, even under Impella support. For example, EDP was higher with the combination of Impella CP at P9 and VA-ECMO at 3000 rpm than with Impella CP at P9 and VA-ECMO at 2000 rpm (15 mmHg vs. 8 mmHg).

**Figure 4:**
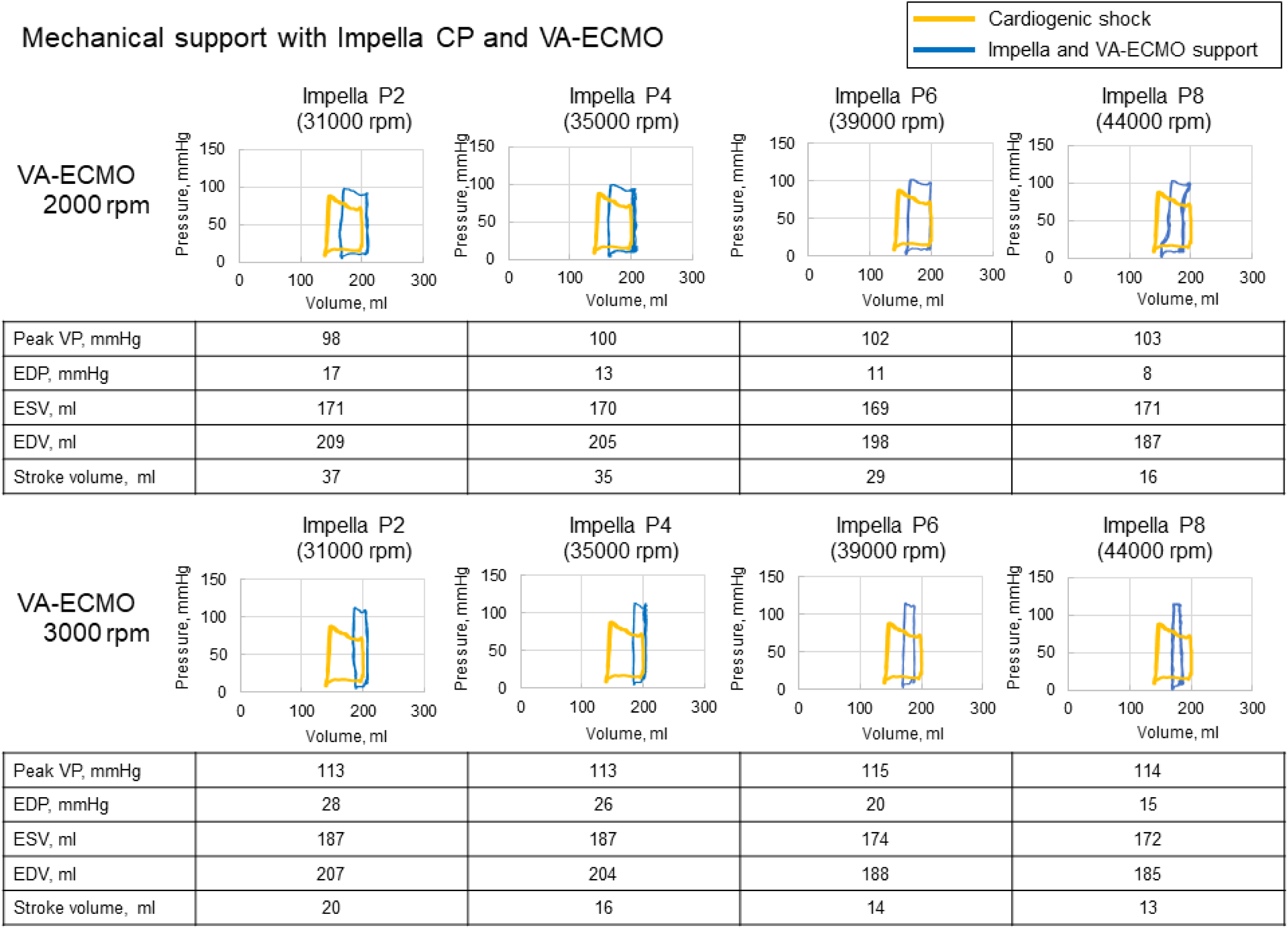
Representative pressure-volume loop of the left ventricle in the cardiogenic shock model with Impella CP and veno-arterial extracorporeal membrane oxygenation (VA-ECMO). EDP, end-diastolic pressure; EDV, end-diastolic volume; ESV, end-systolic volume; LVP, left ventricular pressure. Yellow line, cardiogenic shock; Blue line, Impella and VA-ECMO support.

**Table 3.** Hemodynamics with a combination of Impella CP and VA-ECMO support.

|  |  |  | Impella CP |  |  |  |  |
| --- | --- | --- | --- | --- | --- | --- | --- |
|  |  |  | P0<br>0 rpm | P2<br>31 000<br>rpm | P4<br>35 000<br>rpm | P6<br>39 000<br>rpm | P8<br>44 000<br>rpm |
| VA-ECMO | 0<br>rpm | Mean total<br>flow, L/min | 2.70 | 3.64 | 3.82 | 3.96 | 4.14 |
|  |  | Pressure at<br>A, s/d (m),<br>mmHg | 73/23<br>(46) | 82/48<br>(64) | 84/55<br>(68) | 86/59<br>(71) | 90/63<br>(74) |
|  |  | Mean<br>pressure at<br>VR, mmHg | 15.0 | 11.3 | 10.5 | 10.0 | 9.3 |
|  | 2<br>000<br>rpm | Mean total<br>flow, L/min | 3.99 | 4.26 | 4.37 | 4.54 | 4.71 |
|  |  | Pressure at<br>A, s/d (m),<br>mmHg | 88/61<br>(74) | 92/69<br>(79) | 93/72<br>(81) | 96/79<br>(86) | 98/83<br>(89) |
|  |  | Mean<br>pressure at<br>VR, mmHg | 9.7 | 8.1 | 8.3 | 7.6 | 7.3 |
|  | 3<br>000<br>rpm | Mean total<br>flow, L/min | 4.47 | 4.69 | 4.69 | 4.78 | 5.24 |
|  |  | Pressure at<br>A, s/d (m),<br>mmHg | 106/91<br>(97) | 108/96<br>(101) | 109/97<br>(102) | 110/101<br>(105) | 120/114<br>(116) |
|  |  | Mean<br>pressure at<br>VR, mmHg | 7.0 | 6.1 | 6.3 | 6.0 | 5.0 |

A, aorta; VR, venous reservoir; s, systolic; d, diastolic; m, mean.

The results of the flow at the aorta, iliac arteries, and brachiocephalic artery are presented in Table 4. The mean total flow increased by increasing either the Impella or VA-ECMO support level. The mean aortic flow increased with Impella flow, but decreased with increasing VA-ECMO support. At 3000 rpm of VA-ECMO with Impella from P0 to P4, retrograde flow was observed in the aorta. However, antegrade flow in the aorta was observed at high Impella flow levels (P6 and P8). Importantly, there was a discrepancy in the flow between the aorta and distal artery (iliac arteries). The balance of the support level between Impella and VA-ECMO induces the discrepancy because Impella generates antegrade flow, whereas VA-ECMO generates retrograde flow. The mean flow of the brachiocephalic artery increased by increasing either the Impella or VA-ECMO support level, irrespective of the direction of the aortic flow.

**Table 4.** Mean flow in the aorta, iliac artery, and brachiocephalic trunk.

|  |  |  | Impella CP |  |  |  |  |
| --- | --- | --- | --- | --- | --- | --- | --- |
|  |  |  | P0<br>0 rpm | P2<br>31 000<br>rpm | P4<br>35 000<br>rpm | P6<br>39 000<br>rpm | P8<br>44 000<br>rpm |
| VA-ECMO | 0 rpm | Aorta, L/min | 2.28 | 3.03 | 3.21 | 3.34 | 3.47 |
|  |  | Iliac artery, L/min | 2.33 | 3.14 | 3.30 | 3.41 | 3.57 |
|  |  | Brachiocephalic<br>artery, L/min | 0.38 | 0.50 | 0.53 | 0.55 | 0.57 |
|  |  | Total, L/min | 2.70 | 3.64 | 3.82 | 3.96 | 4.14 |
|  | 2 000<br>rpm | Aorta, L/min | 1.41 | 1.63 | 1.82 | 2.16 | 2.37 |
|  |  | Iliac artery, L/min | 3.47 | 3.67 | 3.77 | 3.92 | 4.05 |
|  |  | Brachiocephalic<br>artery, L/min | 0.52 | 0.59 | 0.60 | 0.62 | 0.67 |
|  |  | Total, L/min | 3.99 | 4.26 | 4.37 | 4.54 | 4.72 |
|  | 3 000<br>rpm | Aorta, L/min | -0.27 | -0.05 | -0.10 | 0.07 | 0.60 |
|  |  | Iliac artery, L/min | 3.85 | 4.00 | 4.00 | 4.08 | 4.46 |
|  |  | Brachiocephalic<br>artery, L/min | 0.63 | 0.69 | 0.69 | 0.70 | 0.77 |
|  |  | Total, L/min | 4.47 | 4.69 | 4.69 | 4.78 | 5.24 |

4 Total flow=flow of the iliac artery plus brachiocephalic artery.

### Self-recirculation with Impella circulatory support device

TARF, including native aortic regurgitant flow with self-recirculation, was observed in both soft and hard silicone valves (Figure 5). The regurgitant fraction in the soft and hard silicone valves without Impella was 14.5% and 21.1%, respectively (Figure 5A and 5B). After implantation of Impella, The regurgitant fraction increased in both soft and hard silicone valves (16.8% and 24.4% at P0, 28.1% and 42.8% at P9, respectively). With the support of Impella, the regurgitant fraction in the hard valve was numerically larger than that in the soft valve.

**Figure 5:**
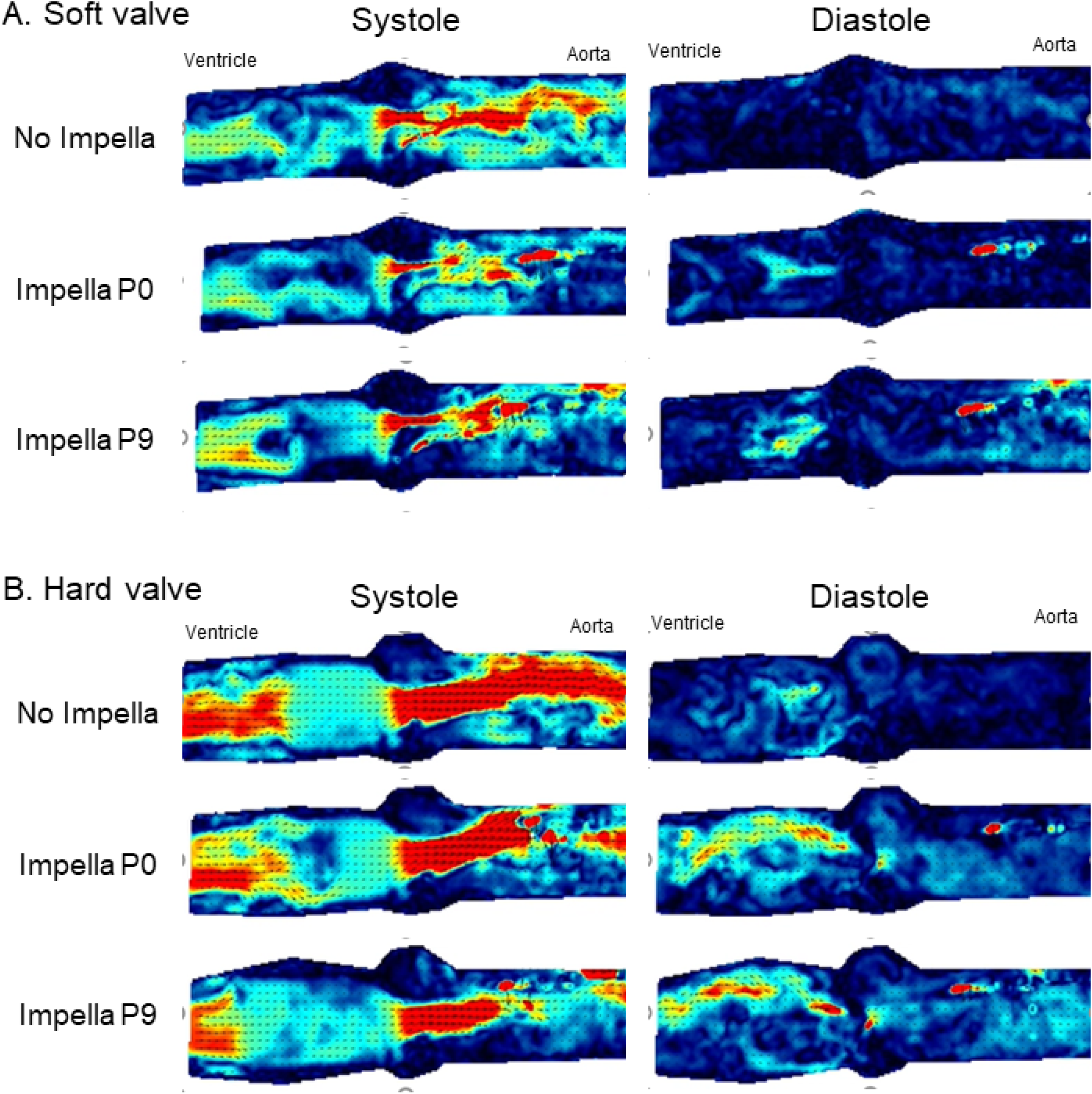
Representative images of particle imaging velocimetry (PIV). (A) Flow distribution with soft valve without and with Impella P0 and P9. (B) Flow distribution with hard valve without and with Impella P0 and P9.

## Discussion

We successfully created an experimental pulsatile flow system with venous function that can be used to assess MCS, including Impella and VA-ECMO. The main findings of the current study are as follows: (1) the Impella circulatory support device could increase forward flow and arterial pressure and decrease EDV and EDP (LV unloading); (2) VA-ECMO could increase arterial pressure owing to increased LV afterload and alter the direction of the aortic flow (antegrade or retrograde) depending on the flow level with VA-ECMO; (3) VA-ECMO could significantly reduce venous pressure (venous unloading) compared with Impella CP; (4) the combination of Impella and VA-ECMO could lead to the greatest hemodynamic support with LV unloading (Figure 6); and (5) Effective Impella flow was reduced during ventricular diastole when aortic insufficiency was present owing to self-recirculation with the Impella device.

**Figure 6:**
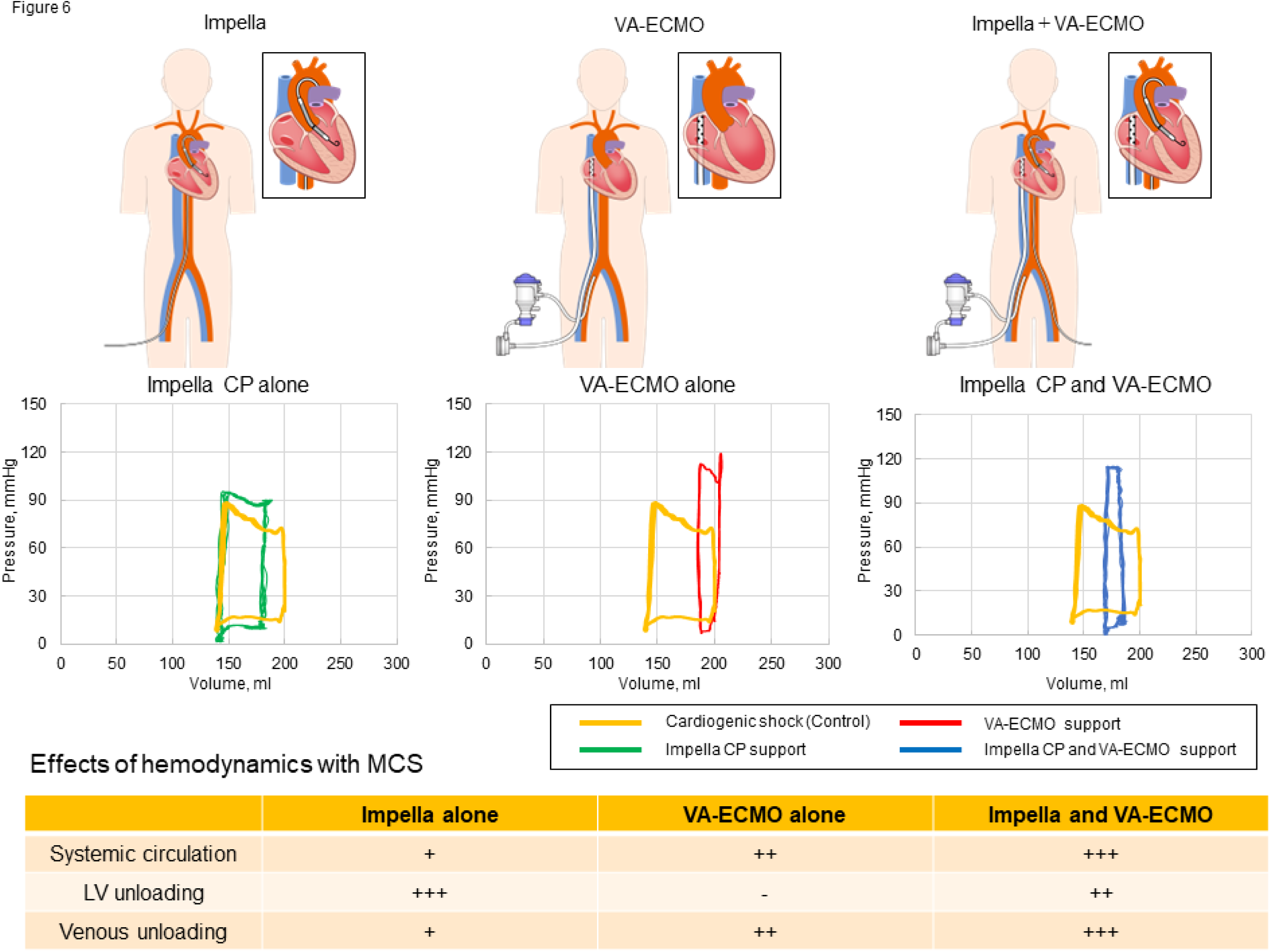
Representative pressure-volume loop of Impella alone, veno-arterial extracorporeal membrane oxygenation (VA-ECMO) alone, and combination of Impella and VA-ECMO and effects of hemodynamics with mechanical circulatory support (MCS).

### Novel pulsatile flow model with venous system

Clinical evidence, including high-quality clinical trials such as randomized controlled studies for patients with CS, is scarce because of the difficulties inherent in these challenging populations. Therefore, in vitro experimental and animal studies are valuable for understanding the hemodynamics of MCS in this field. Several animal studies have been conducted to assess the hemodynamics with Impella. ^23, 24^ However, it is difficult to perform detailed assessments even in animal studies. The pulsatile flow circulation system in this study was created based on our previous model ^16^ and expanded to assess hemodynamics with clinically available MCS devices.

### Hemodynamic support with Impella or VA-ECMO

Several preclinical and clinical studies have demonstrated the advantage of the Impella circulatory support device for high-risk coronary intervention or CS, resulting from increased blood pressure and LV unloading. ^9, 25^ Similar to previous reports, this study showed ventricular unloading with Impella, and significant unloading was observed with increased Impella flow.

The benefit of VA-ECMO is an increase in aortic pressure and a decrease in venous pressure (venous unloading). However, an increase in EDV/EDP could be a problem for early heart recovery, especially after AMI. Increased blood pressure and EDV are associated with a greater pressure-volume area, resulting in greater myocardial oxygen demand, which prevents myocardial healing. ^26^ This study indicated that the effect of decreasing venous pressure (venous unloading) was greater with VA-ECMO than with Impella CP. Previous reports have shown that elevated central venous pressure resulting in worsening renal function leads to cardiovascular events. ^27, 28^ Therefore, VA-ECMO should be an option to maintain hemodynamics in patients with severe CS who have high central venous pressure.

### Combination of Impella and VA-ECMO therapy

Although the combination of Impella and VA-ECMO showed better clinical outcomes than VA-ECMO alone, ^10, 11^ the hemodynamics with the combination therapy remains unknown. In the current study, the combination of Impella and VA-ECMO showed resulted in the greatest hemodynamic support (greatest total flow and arterial pressure). The settings of the flow levels in patients on Impella and VA-ECMO are extremely important for patient survival. In the severe condition of AMI, VA-ECMO might be needed for hemodynamic stability, although VA-ECMO results in delayed heart recovery due to increased LV afterload. Clinical studies including patients with AMI suggested that the incidence of death and rehospitalization with a smaller infarct myocardium was lower than that with a larger infarct myocardium. ^29^ In an animal study, strong Impella support led to smaller myocardial infarction than no support. ^24^ Therefore, increased Impella support leading to lower EDP/EDV should be considered for the recovery of cardiac function once hemodynamics stabilizes.

Most recently, a large registry including patients with CS from the Mayo Clinic showed that the mortality in patients with refractory shock was extremely high. ^12^ Moreover, hypoperfusion leads to greater mortality than hypotension. ^30^ Therefore, early hemodynamic support with sufficient cardiac output is imperative to increase the survival rate. Clinically, insufficient support with Impella due to suction was observed in some cases, even in cases of volume overload due to cardiac failure. Possible reasons for suction were low volume, incorrect pump position, and inadequate LV filling due to right ventricular failure. One of the issues in patients with acute heart failure is hypoxic pulmonary vasoconstriction. ^31^ When Impella support is insufficient, escalation with VA-ECMO or a left ventricular assist device (LVAD) should be considered. Additionally, in cases with precapillary pulmonary hypertension, decreasing pulmonary resistance with a pulmonary vasodilator, such as a phosphodiesterase III inhibitor and inhaled nitric oxide, may be another option to increase cardiac output. ^32^

### Assessment of flow with the combination of Impella and VA-ECMO

North-south syndrome is one of the complications after the implantation of VA-ECMO. Forward flow was observed even in VA-ECMO support when the flow from the proximal aorta was greater than that from the distal aorta, and increased flow with VA-ECMO resulted in retrograde flow at the aorta. The present study showed that retrograde aortic flow was observed when the VA-ECMO support was strong (3000 rpm). In addition, flow stagnation might be present between P4 (35000 rpm) and P6 (39000 rpm) with Impella CP under VA-ECMO (3000 rpm), because the flow of the aorta was almost zero. A recent case report described the presence of a large mobile thrombus in the descending aorta in a patient with CS supported by a combination of Impella (2.0 L/min) and VA-ECMO (3.0 L/min). ^33^ Theoretically, when the retrograde flow of VA-ECMO is equivalent to the forward flow, the aortic flow is likely to stagnate. From the present study and case report, both Impella and VA-ECMO with high-flow support in patients with CS might be at risk of inducing thrombus formation because of the stagnation of the aortic flow, although the situation should be different based on cardiac function.

### Self-recirculation with Impella circulatory support device

Impella is a catheter with a small built-in axial flow pump that pulls blood from the LV through the inlet area and expels blood from the catheter into the ascending aorta. To the best of our knowledge, flow analysis using PIV with Impella in pulsatile circulatory systems has not been reported. The present study identified the occurrence of self-recirculation during valve closing in the presence of aortic insufficiency. Moreover, the amount of self-recirculation increased with an increase in the Impella flow level when an aortic valve inducing aortic insufficiency was used. Notably, the effect of LV unloading with Impella is diminished by self-recirculation in patients with significant aortic insufficiency.

Recirculation was previously reported in patients with aortic insufficiency after LVAD implantation. ^34^ The blood delivered into the aorta by the LVAD recirculates back into the LV instead of reaching systemic circulation. Clinically, a patient with severe aortic insufficiency following LVAD implantation may present with high LVAD flow and increased afterload, which has been reported as a major limitation following LVAD implantation. ^35^ Therefore, current guidelines indicate that significant aortic insufficiency is considered a contraindication in LVAD support. ^35, 36^

The Impella circulatory support device is placed through the aortic valve; therefore, aortic insufficiency may be observed due to the loss of coaptation between the aortic valve leaflets. Currently, Impella 2.5/CP, which can be percutaneously inserted, is often used to treat AMI complicated by CS. When the escalation of the mechanical circulatory device is required for refractory CS, Impella 5.0 is one of the options for escalation. However, in patients with significant aortic insufficiency, VA-ECMO or LVAD implantation should be considered instead of Impella 5.0 implantation, as Impella support may not be effective in recovering hemodynamics. Therefore, attention should be paid to the possibility of insufficient hemodynamic support in patients with significant aortic insufficiency.

### Significance of clinically relevant in vitro circulation model

Currently, MCS devices are widely used in patients with CS. However, obtaining strong evidence in this field is difficult. For example, the recommendation of IABP use has been downgraded, and there has been no randomized controlled study of VA-ECMO. Thus, in vitro clinically relevant studies with circulation models are imperative for assessing the influence of MCS on hemodynamics in CS patient analogs with various settings and answering clinical questions in real-world clinical practice. We believe that the present circulation system should be useful for hemodynamic assessment, even for other MCS.

This study has several limitations. First, the present study was conducted using an in vitro circuit without the right ventricle. Therefore, the hemodynamics of the right heart could not be assessed. Second, we assessed only one setting of CS status using Impella and VA-ECMO. Clinical situations include many different conditions; therefore, the data from the present study may not be widely applied to clinical situations. However, the trend of MCS effects can be understood. Third, the present study suggests that the combined use of Impella and VA-ECMO might be the best strategy with strong hemodynamic support for patients with severe CS, but higher incidence of complications, including cannula-site bleeding, following Impella implantation has been reported than with IABP. Therefore, careful assessment of the bleeding risk is required for the treatment with MCS. Fourth, recirculation was assessed using a silicone valve; therefore, the coaptation of the valve might be different from that of the native human valve. Nevertheless, the present findings may help understand the mechanisms of aortic regurgitant flow under Impella support.

In conclusion, Ventricle unloading is a specific advantage of the Impella circulatory support system that leads to improved hemodynamics. VA-ECMO is a powerful device to support hemodynamics with venous unloading, although an increased cardiac load was induced. The combination of Impella and VA-ECMO should be a valuable option for patients with severe CS. Self-recirculation with Impella reduces hemodynamic support in patients with aortic insufficiency. We believe that the findings using the present pulsatile circulation system may provide valuable insights into optimizing the MCS strategy for the management of patients with CS.

## Funding

This study was supported by AMED under Grant Number JP22mk0101179 and in part by the Subsidy Program for the Development of International Standards for the Evaluation of Innovative Medical Devices and Regenerative Medicine Products of the Ministry of Health, Labor and Welfare, Japan. The Impella circulatory support system and VA-ECMO were provided by Abiomed Japan and Senko Medical Instruments.

## Conflict of Interest

Dr. Yahagi received remuneration from Abiomed Japan. All other authors have no relationships relevant to the contents of this article to disclose.

## Abbreviations

CS: Cardiogenic shock
MCS: Mechanical circulatory support
IABP: intra-aortic balloon pumping
VA-ECMO: veno-arterial extracorporeal membrane oxygenation
AMI: acute myocardial infarction
LV: left ventricular
Nd:YAG: neodymium-doped yttrium aluminum garnet
TARF: total aortic regurgitant flow
RF: regurgitant fraction
PIV: particle imaging velocimetry
Ees-end-systolic LV elastance
Ea: arterial elastance
LVAD: left ventricular assist device.

## Data Availability

We included a statement in the Methods.

## Notes

### Competing Interest Statement

The authors have declared no competing interest.

### Author Declarations

IRB review has not been conducted because this study was performed using a pulsatile flow model.

